# Introduction and transmission of SARS-CoV-2 B.1.1.7 in Denmark

**DOI:** 10.1101/2021.06.04.21258333

**Authors:** Thomas Y. Michaelsen, Marc Bennedbæk, Lasse E. Christiansen, Mia S. F. Jørgensen, Camilla H. Møller, Emil A. Sørensen, Simon Knutsson, Jakob Brandt, Thomas B. N. Jensen, Clarisse Chiche-Lapierre, Emilio F. Collados, Trine Sørensen, Celine Petersen, Vang Le-Quy, Mantas Sereika, Frederik T. Hansen, Morten Rasmussen, Jannik Fonager, Søren M. Karst, Rasmus L. Marvig, Marc Stegger, Raphael N. Sieber, Robert Skov, Rebecca Legarth, Tyra G. Krause, Anders Fomsgaard, The Danish Covid-19 Genome Consortium (DCGC), Mads Albertsen

**Affiliations:** Department of Chemistry and Bioscience, Aalborg University; Aalborg, Denmark; Centre of Excellence for Health, Immunity and Infection (CHIP), Department of Infectious Diseases, Rigshospitalet, University of Copenhagen; Copenhagen, Denmark; Department of Applied Mathematics and Computer Science, Technical University of Denmark; Lyngby, Denmark; Infectious Disease Epidemiology & Prevention, Statens Serum Institut; Copenhagen, Denmark; Infectious Disease Preparedness, Statens Serum Institut; Copenhagen, Denmark; Unit for Research Data Services (CLAAUDIA), Aalborg University; Aalborg, Denmark; Department of Virus & Microbiological Special Diagnostics, Statens Serum Institut; Copenhagen, Denmark; Center for Genomic Medicine, Rigshospitalet; Copenhagen, Denmark; Department of Bacteria, Parasites and Fungi, Statens Serum Institut; Copenhagen, Denmark

## Abstract

In early 2021, the SARS-CoV-2 lineage B.1.1.7 became dominant across large parts of the world. In Denmark, comprehensive and real-time test, contact-tracing, and sequencing efforts were applied to sustain epidemic control. Here, we use these data to investigate the transmissibility, introduction, and onward transmission of B.1.1.7 in Denmark. In a period with stable restrictions, we estimated an increased B.1.1.7 transmissibility of 58% (95% CI: [56%,60%]) relative to other lineages. Epidemiological and phylogenetic analyses revealed that 37% of B.1.1.7 cases were related to the initial introduction in November 2020. Continuous introductions contributed substantially to case numbers, highlighting the benefit of balanced travel restrictions and self-isolation procedures coupled with comprehensive surveillance efforts, to sustain epidemic control in the face of emerging variants.

---

A year into the SARS-CoV-2 pandemic it became clear that variants with increased transmissibility and/or reduced vaccine efficacy would have a profound impact on our lives, including prolongation of the pandemic as well as increasing morbidity and mortality. On 14 December 2020, Denmark received a notification from the Early Warning and Response System of the European Union (EWRS) about a novel SARS-CoV-2 lineage (B.1.1.7) spreading fast throughout the United Kingdom (UK). The notification was a direct response to genomic sequences uploaded to the GISAID database by the Danish Covid-19 Genome Consortium (DCGC). On 18 December 2020, the New and Emerging Respiratory Virus Threats Advisory Group (NERVTAG), advisers to the British Health Authorities, released a public statement, reporting a 71% (95% CI: [67%,75%]) increased transmissibility for the B.1.1.7 lineage and elevated it to variant-of-concern (VOC) (*1*). The B.1.1.7 lineage appeared in the UK in September 2020 and has since spread across the world (*2*), including Denmark where the first identified case was sampled on 14 November 2020 (*3*). Several studies have since estimated an increased transmissibility of B.1.1.7, ranging from 29% to 90% (*4–7*). This is in line with early estimates of 36%-55% from Denmark (*8, 9*). Hypotheses for increased transmissibility include higher viral load, longer infection period, and better receptor binding (*10–12*).

The B.1.1.7 lineage contains several genetic changes compared to the specimens first identified in Wuhan, China. Most notably is the spike protein amino acid substitution N501Y which enhances binding to the ACE2 receptor and is thought to increase transmissibility (*12*). Acquired mutations can be used to construct the phylogenetic relationship between genomes and by coupling with temporal information, transmission timing and dynamics of emerging lineages can be inferred (*13*). These approaches have been used by several studies to gain insights into the SARS-CoV-2 pandemic on a national level (*6, 7, 14–17*).

Due to the early warning from the UK and an already extensive sequencing effort in Denmark, it was possible to predict growth dynamics for B.1.1.7 in Denmark, when it was still less than 2% of all circulating SARS-CoV-2 lineages. Hence, on 2 January 2021, B.1.1.7 was predicted to exceed 50% relative abundance nationwide in the middle of February (*3, 18*), which formed a key argument in the political decision to further extend the lockdown at the time and intensify contact tracing on all B.1.1.7 cases (*19*). In this study we substantiate early results by providing an updated estimate of increased transmissibility of B.1.1.7. Furthermore, using the detailed and comprehensive nationwide data, we outline the dynamics of B.1.1.7 in Denmark to evaluate the impact of introductions and regional restrictions on the prevalence and spread of B.1.1.7.

We modelled the growth rate of B.1.1.7 relative to other lineages in the period from 4 January to 7 February 2021. The study period was chosen as both the PCR testing and sequencing capacity increased substantially until January, reaching a weekly rate of >13,000 tests pr. 100,000 people and >75% of all positive samples having a high-quality genome (fig. 1A, fig. S1). Furthermore, a window with stable restrictions was enforced from January 4th until partial reopening of primary schools from February 8th. A Poisson regression model was fitted on daily counts of B.1.1.7 to estimate increased transmissibility of B.1.1.7 relative to all other lineages. We found a growth rate of 0.021 and -0.062 per day for B.1.1.7 and other lineages respectively, corresponding to an increased transmissibility of 58% (95% CI: [56%,60%]) for B.1.1.7 assuming a generation time of 5.5 days (*10, 20*). We did not observe significant differences in transmissibility between regions 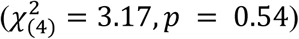.

**Fig. 1.**
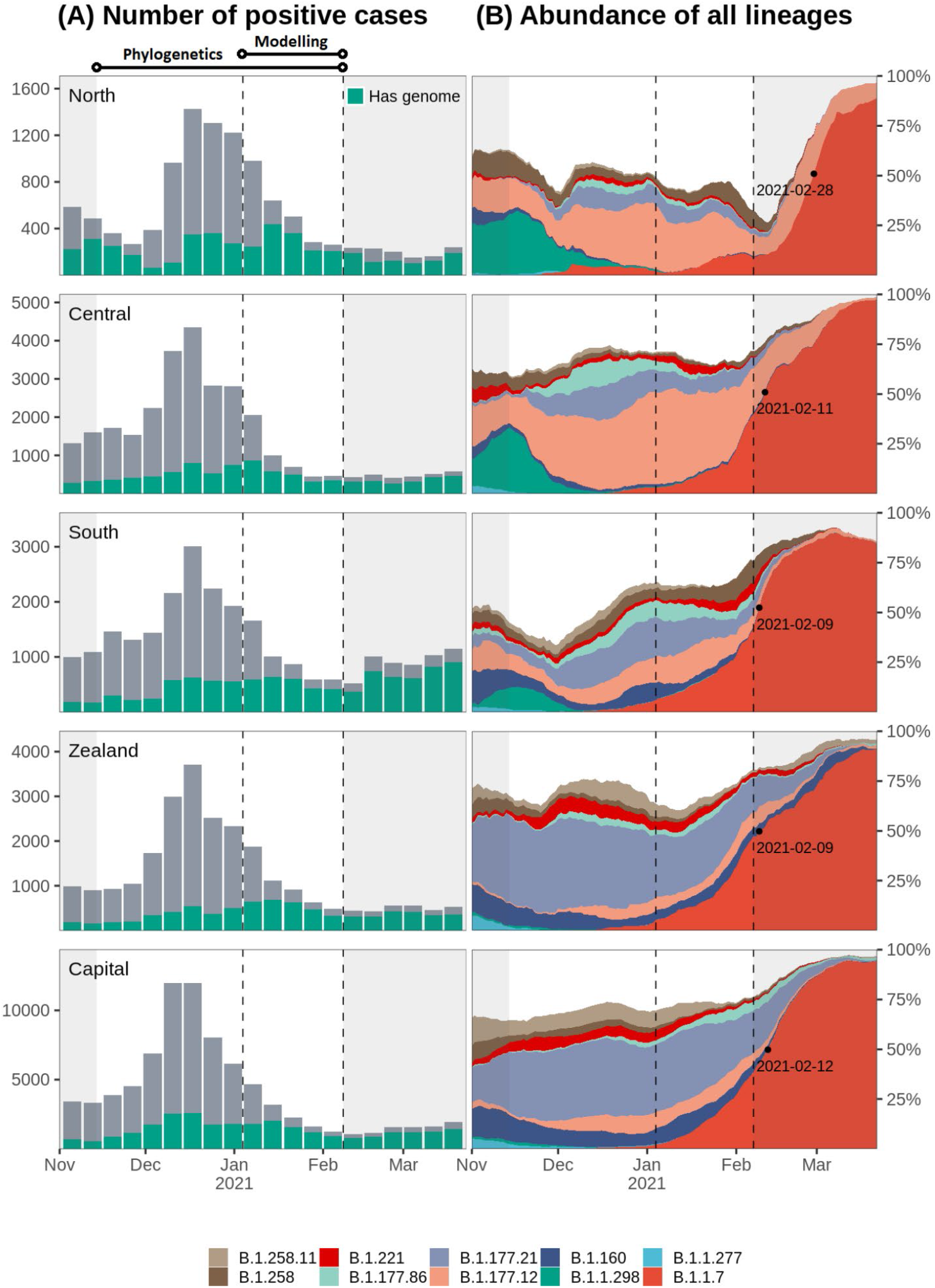
Sequencing rate and lineage dynamics. Each row corresponds to the region highlighted to the far left. The two vertical dashed lines indicate the beginning and end of study period used to infer B.1.1.7 transmissibility, while the non-shaded area shows the period used for phylogenetic analysis. The time outside the study are shaded in grey. **(A)** sequencing rate across time. Total height corresponds to the total number of cases for each week. Green bars indicate the number of cases for which a high-quality genome was available (less than 3000 ambiguous bases). **(B)** relative abundance of ten most abundant PANGO lineages across Denmark in the study period. Computed from rolling averages of daily cases with genome available over a 14-days window. Less abundant lineages are not shown but included in the unfilled area of the plots. The black dot and date corresponds to the point where B.1.1.7 crosses 50% relative abundance.

After the study period ended, case numbers of B.1.1.7 in South Denmark and Capital increased to higher levels than what was predicted by our model (fig. S2A), likely caused by large superspreading events, evident from the rapid increase of unique local haplotypes in South Denmark (H17 and H87) and Capital (H43) regions (fig. S2B). All regions crossed 50% relative abundance of B.1.1.7 with a few days difference (fig. 1B), except North Denmark, with 1-2 weeks delay and several incidences of B.1.1.7 decline (fig. 1B).

Introductions into Denmark and onward transmission between regions were investigated using a time-scaled phylogenetic tree of 1976 Danish B.1.1.7 genomes and 3611 representative international sequences from outside Denmark (fig 2A). We used pastML (*21*) to perform an ancestral state reconstruction, to infer if sequences were introduced from outside Denmark or transmitted from another Danish region. We define an introduction lineage as a phylogenetic group of Danish sequences that share the same introduction event, similar to the definition of UK transmission lineages in (*15*). We do not infer the timing of introduction events from the phylogeny (*7, 15, 22*), but instead use the first occurrence of an introduction lineage as a proxy justified by the intense testing and high sequencing rate (fig. S1). An introduction lineage can be further subdivided into transmission clusters from the ancestral state reconstruction, which is based on the regional and temporal sub-distributions of an introduction lineage. The inferred connections between transmission clusters outlines the direction of transmission between regions. The first cases of B.1.1.7 observed in the Capital region of Denmark on 14 November 2020 defines the introduction lineage CA1, which contained the majority of Danish sequences (736 of 1976, 37%) across all Danish regions (fig. 2B, fig. S3). We tried to resolve the origin of CA1, but detailed epidemiological investigations revealed no direct or indirect travel link and the ancestral state could not be resolved with good confidence. The CA1 introduction lineage is associated with the first reported cases of B.1.1.7 in all Danish regions, including the first North Denmark transmission cluster CA1-135. This transmission cluster was part of a large but local outbreak verified by epidemiological investigations and primarily associated with a single local superspreading event (haplotype H77, fig. S2B). CA1-135 dominated the North Denmark cases until it was last observed in week 2 (fig. 2B). This contrasts the course of the first transmission clusters in other regions, which showed similar initial growth (CA1-108 in South Denmark and CA1-124 in Zealand) but remained at high prevalence throughout the study period.

**Fig. 2.**
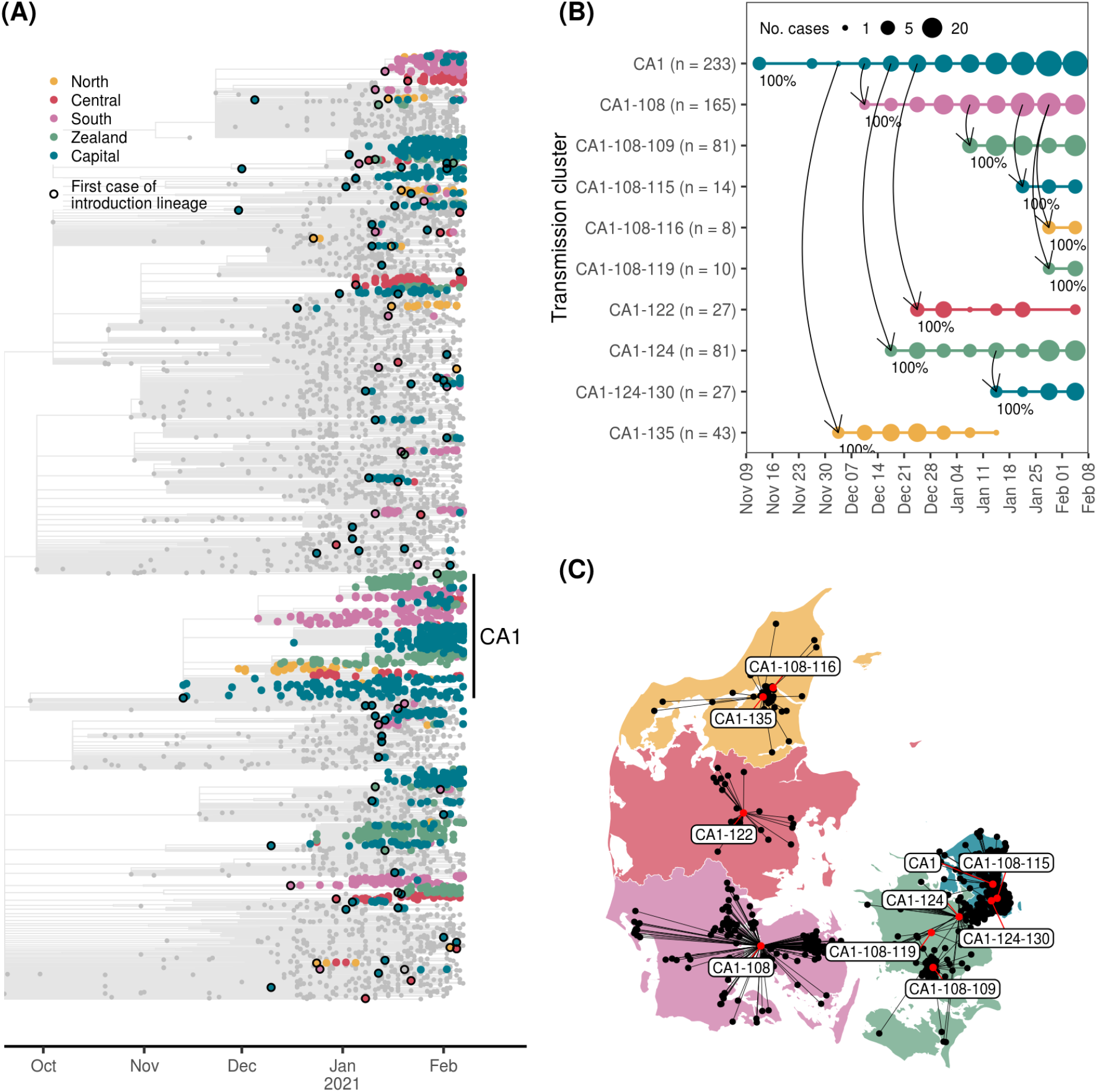
Early introduction of B.1.1.7 and onwards transmission between Danish regions. (**A**) time-adjusted phylogenetic tree. Tree branches and non-Danish sequences are coloured grey. First sequence in a transmission lineage is used to infer time of importation and highlighted with a black circle. (**B**) and (**C**) shows a focused temporal analysis of the CA1 introduction lineage, focusing on descending clusters with more than 5 cases. (**B**) the temporal span and dynamics of onwards transmission across regions. Each descendent transmission cluster of CA1 is indicated with a solid horizontal line, with points at each week sized according to the number of cases. The vertical curved arrows indicate direction of transmission. The marginal probability of the observed transmission link is indicated at each arrow. In (**C**) the corresponding cases are placed on a map of Denmark using black points, with up to 5 km of random jitter added. Cases from the same transmission cluster are linked by black lines to their centroid location indicated by a red dot. Labels indicate cluster id’s.

Interestingly, all transmission clusters appearing up until week 1 in North Denmark were not observed in the remaining study period, which were not true for other regions where a constant level of sustained transmission was present for many larger transmission clusters (fig. 3A). In North Denmark, 7 of 11 municipalities had elevated restrictive measures until 16 November 2020 due to circulation of mink-related lineages (*23*). The intensified contact-tracing and genomic surveillance in that period likely kept the reproductive numbers for North Denmark below that of other Danish regions, making it difficult for any cluster to develop from local to sustained transmission. B.1.1.7 was only designated as VOC from late December 2020 onwards (*1*) and authorities were not able to do sufficient genome-based contact tracing due to low number of samples sequenced (around 25% of positive cases) and slow turnaround (median 11 days) at that time. Therefore, the dynamics in North Denmark is likely not attributed to focused contact tracing of B.1.1.7. This contrasts the current situation in June 2021, with substantially improved sequencing capacity (>90% of positive cases and median 5 days turnaround) and rapid mutation of concern screening to initiate contact tracing (*24, 25*).

**Fig. 3.**
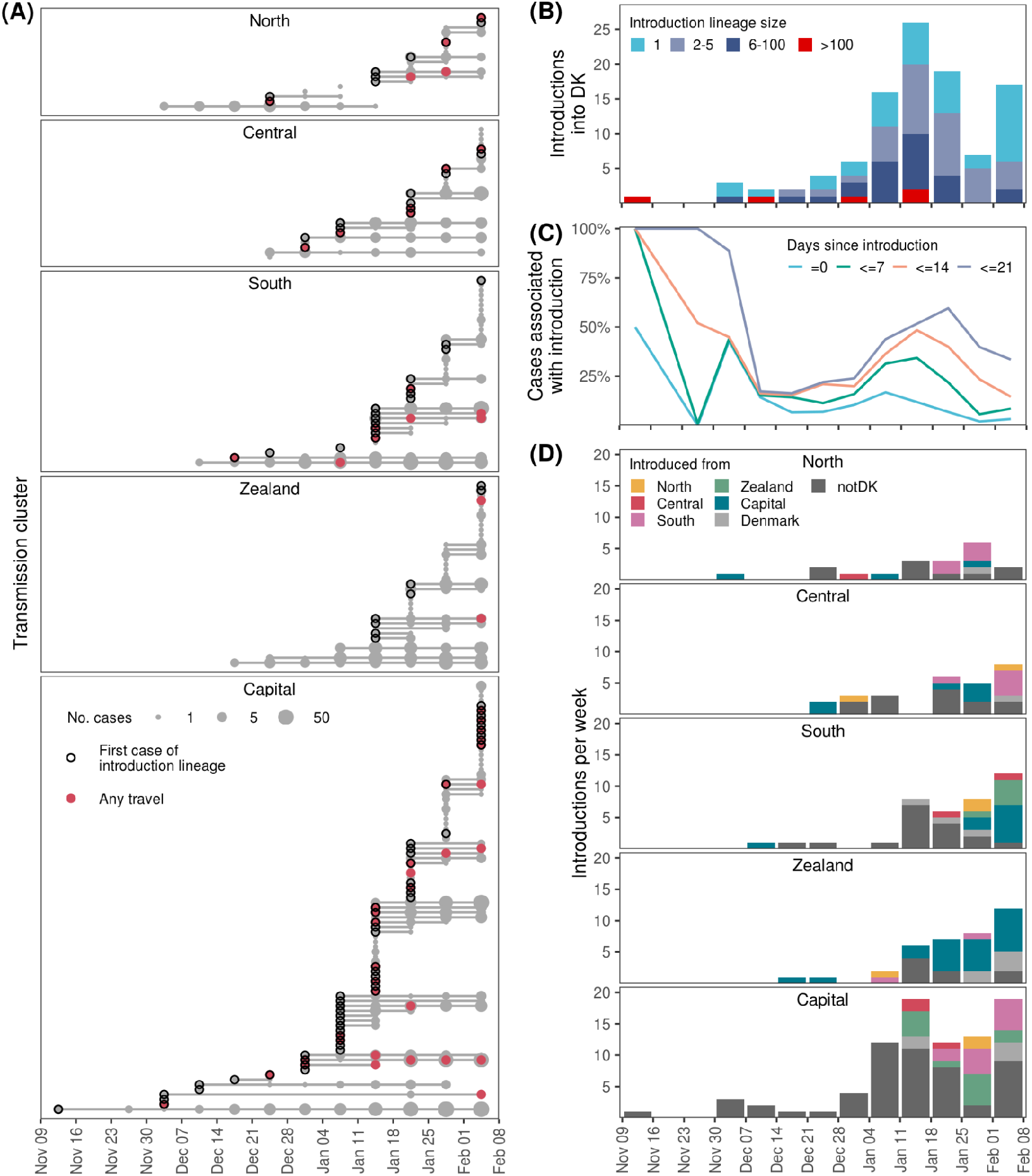
Analysis of importation events into Denmark and onwards transmission between regions. (**A**) the temporal span and development of each transmission cluster, grouped by Danish region. Each transmission cluster is indicated with a solid horizontal line, with points at each week sized according to the number of cases. Circles indicate first observed case of an introduction lineage imported from outside Denmark. Red points indicate presence of cases with travel history in the given week. (**B**) the number of introductions into Denmark across time. Colours indicate the total number of offspring cases associated with each introduction lineage. (**C**) the relative contribution of introductions versus ongoing existing transmission. Introduction-releated cases were defined with various cutoffs for the maximum number of days between the case and the first observed case for each introduction lineage, as indicated by different colours. (**D**) the origin of introductions across time for each region. If there was equal support for multiple regions as origin for an introduction Denmark was used as the origin. Label notDK indicates an introduction from outside Denmark. Only ancestral state changes with a marginal probability >95% were included in the analysis.

Overall, we identified 103 introductions and 107 transmission events between regions of B.1.1.7 in the study period (fig. 3). The majority of introductions occurred during the first half of January 2021 (fig. 3B); 37 introductions showed no onward transmission, 36 introductions had between 2-5 cases of whom 8 exclusively had transmission within the same household, and only 5 caused >100 cases. To investigate the impact of introductions across time we considered early cases of the same introduction lineage to be introduction-related, while later cases occur due to sustained transmission in society. To evaluate this, we incremented the number of follow-up days for cases to be assigned as introduction-related (fig. 3C). Overall, the relative contribution of introduction-related cases peaked during onset of B.1.1.7 and again in mid-January (fig. 3C). Introductions alone (follow-up =0 days in fig. 3C) accounted for around 5%-10% of all cases with substantial temporal variation. This is comparable to a US study on data from the first half of 2020, where travel restrictions also applied (*22*). Using 14 days follow-up since the first occurrence of an introduction to define introduction-related cases within the same introduction lineage, there were periods where >50% of cases were due to importation from outside Denmark (fig. 3C) despite travel-restrictions and self-isolation recommendations being in place.

In collaboration with the Danish Patient Safety Authority which administers the contact tracing efforts in Denmark, we performed detailed epidemiological investigations of registry data to identify direct or indirect travel links for all 1976 cases in this study. A total of 62 persons infected with B.1.1.7 (3%) had recorded travel history to 23 different destinations. The most frequent destinations were the United Arab Emirates (n = 21), United Kingdom (n = 5), and Pakistan (n = 4). We were able to associate 32 (31%) introductions directly to travelling (fig. 3A, fig. S5). We performed a separate ancestral state reconstruction with country-level resolution on international sequences, allowing us to compare the phylogenetically inferred origin to the travel history. Of 33 introductions with travel history, 9 (27%) matched by country and 12 (36%) by high-level geographical region. This highlights the limitations of available genomic data, which is still heavily biased both geographically and temporally (*26*), making high-resolution inference of introduction origin from phylogenetic data alone very difficult. From the ancestral state reconstruction, we inferred the temporal dynamics of B.1.1.7 dispersion between the five major administrative regions of Denmark. The majority of transmission was local and occurred within regions, but would to a large extent cease if not fueled by introductions from other regions or outside Denmark (fig. 3D). The Capital and South Denmark regions with international borders and substantial international travel-activity also received the most introductions, possibly a combined effect of multiple factors such as leisure travel and commuting for work. Interestingly, the Capital region also had the highest number of outgoing transmissions to other regions, acting as an important source for sustaining B.1.1.7 in Denmark, particularly in the Zealand region which has a large number of residents commuting to the Capital region on a regular basis. In many countries the takeover of B.1.1.7 has prolonged or induced further restrictions to sustain epidemic control. In Denmark, genomic surveillance has been key to rapidly predict growth of B.1.1.7, leading to enforced restrictions in effect from 5 January 2021 when B.1.1.7 was still below 2% relative abundance of all SARS-CoV-2 variants (*27*). The enforced restrictions practically eliminated all other circulating variants within 2 months (fig. 1B) and enabled epidemic control to be maintained as society was gradually reopened from 8 February 2021. We validated the initially predicted increased transmissibility of B.1.1.7 and found that superspreading events, identified through sequencing data, could explain deviations from the model (fig. S2). At the time when B.1.1.7 was recognized as VOC, the initial CA1 introduction lineage had expanded considerably across all regions, which substantially accelerated the initial phase of the B.1.1.7 pandemic in Denmark. However, ongoing introductions and regional transmissions continuously fueled and further accelerated growth of this lineage. Since 22 December 2020, authorities enforced elevated restrictions for travelers from the UK to halt B.1.1.7 expansion (*19*), similar to the restrictions enforced for inbound travelers from Italy and Austria in the spring of 2020 to stop SARS-CoV-2 from spreading to Denmark (*28*). Despite these restrictions, our study shows a substantial impact of introductions, suggesting that country-specific restrictions and non-enforced recommendations of self-isolation might be insufficient. Finally, VOC designation of B.1.1.7 and upscaling of capacity in Denmark was too late to hinder the takeover of this lineage, which had to be managed by broad societal restrictions. However, real-time testing, genomic tracking and contact-tracing is now at a level in Denmark which enables control of emerging VOCs like B.1.617.2 (*3*) and allow policy makers to make informed decisions of opening the society while maintaining control of the epidemic in the face of emerging variants.

## Data Availability

This study was conducted on administrative register data. According to Danish law, informed consent is not needed for such research as the publication only contains aggregated results and no personal data is shown. The epidemiological data used in this work is person-sensitive and we are prohibited to make it publicly available according to Danish legislation. SARS-CoV-2 consensus genome sequences associated with this work have been uploaded to the GISAID database in accordance with Danish law (no. 285 vers. 2021-02-27). As consequence, dates are binned by week and maximum spatial resolution is at regional level. Accession numbers are available at https://github.com/TYMichaelsen/B117-DK-introduction. All code used to run the phylogenetic analysis, Poisson regression models, and generate the visualizations used in this work is available at https://github.com/TYMichaelsen/B117-DK-introduction.

## Acknowledgments

We gratefully acknowledge the Danish hospitals for local sequencing and data sharing. Thanks to all data contributors from outside Denmark, including the authors and their originating laboratories responsible for obtaining the specimens, as well as their submitting laboratories for generating the genetic sequence and metadata and sharing it via the GISAID Initiative. We would like to acknowledge the people at Data Integration and Analysis (DIAS), Statens Serum Institute, for managing and maintaining the national MiBa database which much of this work is based on. Thanks to the people at the Danish Patient Safety Authority for providing us with detailed travel records of SARS-CoV-2 positive cases. Thanks to Anna Zhukova, Institut Pasteur for modifying the pastML software to better fit our needs upon request. Finally, a sincere thanks to Edyth Parker, Scripps Research, for insightful phylogenetic discussions.

## Funding

VILLUM FONDEN grant (15510) (MA), Poul Due Jensen Foundation (Corona Danica) (MA), Styrelsen for Forskning og Uddannelse (0238-00002B) (MA)

## Author contributions

TYM analyzed all phylogenetic and epidemiological data and prepared all figures. TYM and MA wrote the paper. TYM, MB, MR, JF, and RLM designed the phylogenetic analysis. LEC and CHM designed the Poisson regression models. MSFJ, RL, and TYM performed epidemiological investigations. EAS, SK, JB, SMK, TBNJ, CCL, EFC, TS, CP, VL, MR, JF, MSe, FTH, MSt, RNS, TYM and MA generated the sequencing data. MA, RS, TGK, AF oversaw the study. All authors reviewed and edited the manuscript.

## Competing interests

All authors declare no competing interests.

## Data and materials availability

This study was conducted on administrative register data. According to Danish law, informed consent is not needed for such research as the publication only contains aggregated results and no personal data is shown. The epidemiological data used in this work is person-sensitive and we are prohibited to make it publicly available according to Danish legislation. SARS-CoV-2 consensus genome sequences associated with this work have been uploaded to the GISAID database in accordance with Danish law (no. 285 vers. 2021-02-27). As consequence, dates are binned by week and maximum spatial resolution is at regional level. Accession numbers are available in the Supplementary Materials (data file S1). All code used to run the phylogenetic analysis, Poisson regression models, and generate the visualizations used in this work is available at https://github.com/TYMichaelsen/B117-DK-introduction.

## Supplementary Materials

Materials and Methods

List of Danish Covid-19 Genome Consortium members

Figs. S1 to S4

Data file S1

References (12, 21, 29-46)

## Notes

### Competing Interest Statement

The authors have declared no competing interest.

### Author Declarations

This study was conducted on administrative register data. According to Danish law, ethics approval is not needed for such research. All data management and analyses were carried out on the Danish Health Data Authority's servers with restricted access. The publication only contains aggregated results and no personal data. The publication is, therefore, not covered by the European General Data Protection Regulation.

