## Supplementary Materials for "Introduction and transmission of SARS-CoV-2 B.1.1.7 in Denmark"

**This PDF file includes:**

Materials and Methods
List of Danish Covid-19 Genome Consortium members
Figs. S1 to S4

**Other Supplementary Materials for this manuscript include the following:**

Data file S1. GISAID acknowledgement table. (.tsv)

### Materials and Methods

Ethics statement

This study was conducted on administrative register data. According to Danish law, ethics approval is not needed for such research. All data management and analyses were carried out on the Danish Health Data Authority's servers with restricted access. The publication only contains aggregated results and no personal data. The publication is, therefore, not covered by the European General Data Protection Regulation.

Study data

We used comprehensive register data from all persons with a positive SARS-CoV-2 RT-PCR test in Denmark. Detailed epidemiological records were retrieved from the Danish Patient Safety Authority. We used the Danish civil registration number, which is a unique personal identifier, to link the two datasets. Daily numbers of RT-PCR tested individuals were obtained from (*29*). We restricted the study period to include the first observation of B.1.1.7 in Denmark on 14 November 2020 (week 46) until 7 February 2021 (week 5) which marked the reopening of Danish primary schools. To estimate transmissibility, we focused on a period with stable restrictions from 4 January (week 1) to 7 February, 2021 (week 5).

Sequencing data

Throughout the study period, a subset of positive RT-PCR tests were selected for sequencing by The Danish COVID-19 Genome Consortium (DCGC), established in March 2020 to assist public health authorities by providing rapid genomic monitoring of the spread of SARS-CoV-2. Selection was done by Ct-values, using cutoffs between 30-38 in the study period (*30*). Whole genome amplification of SARS-CoV-2 was performed using a modified version of the ARTIC tiled PCR scheme (*31*) targeting a total 33 overlapping amplicons between 1000-1500 bp, and a custom 2-step PCR strategy was used for barcoding the amplicon libraries. Barcoded libraries were normalized and pooled and prepared for sequencing with the SQK-LSK109 ligation kit (Oxford Nanopore) and sequencing was performed on the MinION device using R.9.4.1 flowcells (Oxford Nanopore). A complete protocol can be found at protocols.io (*32*). The raw sequencing data was basecalled using Guppy v.3.6.1 and demultiplexed using a custom cutadapt v.2.10 (*33*) wrapper. Generation of consensus sequence was done using the function "artic minion" with default settings from the Arctic Network bioinformatics protocol v.1.1.0 (*34*), which uses Medaka v.1.0.3 for consensus calling. The consensus sequences were masked in the beginning (54 bases) and end (67 bases) to avoid primer biases. A consensus sequence with less than 3000 ambiguous bases (approx. 10%) were considered a "high-quality genome" and used in the national SARS-CoV-2 surveillance. If not, the consensus sequence was considered failed and not used.

Calculating transmissibility

We modelled the daily counts of B.1.1.7 and all other variants using Poisson regression:

$$P_t = I_t T_t^{\beta} S_t$$

Here $P_t$ is the expected daily counts, $T_t$ is the daily number of tested, $S_t$ is the proportion of positive tests with a genome. $I_t$ is a measure of incidence depending on the location (danish region), time (date) and lineage (either B.1.1.7 or any other lineage). $S_t$ and $T_t$ are given as input. As activity in society fluctuates systematically over time this leads to autocorrelation in the observed number of cases. We incorporated an AR1 autocorrelation structure of date for each region separately, using the glmmTMB R-package (*35*). Using R notation this leads to the following formula specification:

$$Pt \sim Region + Date + type + Region{:}type + type{:}Date + log(Tt) + offset(log(St)) + ar1(Date + 0|Region)$$

Where Type indicates either B.1.1.7 or all other lineages. There are interactions between Type and Region and Date to allow for different prevalence for the different types in the different regions and different growth rates, respectively. $\beta$ is estimated by adding $log(T_t)$ and $log(S_t)$ is included as an offset - the logarithms are due to using log as the natural link function in the generalized linear model. We also explored alternative relevant models and compared them to our chosen model using the Akaike Information Criterion (AIC). As we observed a substantial difference in the development of B.1.1.7 in North region, we tested if a region-specific estimate of growth rate could be justified by the data. The AIC increased by 6 (2306.9 to 2312.9) and none of the region-specific growth rates were significant (all $p > 0.25$). We also verified that the autocorrelation structure was needed, as AIC increased substantially by 112 (2306.9 to 2418.9) if the AR1 was dropped from the model.

Phylogenetic analysis

Consensus sequences from DCGC were collected until 17 April 17 2021 and global reference consensus sequences from GISAID were downloaded 15 April 2021. These were labeled with phylogenetic clades and lineages with Pangolin v2.2.2 (*36*) using the PANGO nomenclature (*37*). The data was subset to B.1.1.7 only. The global reference data was then downsampled at random to maximum 20 sequences per collection week and country. Sequences were aligned to the Wuhan-Hu-1 MN908947.3 reference using mafft v7.471 (*38*). We then constructed an initial maximum likelihood tree using IQ-tree v2.0.3 using a general time reversible model (*39*) with empirical base frequencies and a free-rate model (*40*) with three categories. This model was predicted as the best model for maximum likelihood phylogenetic construction assessed using ModelFinder (*41*). From the initial tree, sequences with >3 times residuals from the interquartile range in a root-to-tip regression (*42*) and sequences with branch lengths longer than the 99% percentile of the branch length distribution were discarded. From the filtered sequences we ran 1000 ultrafast bootstrap (UFboot, (*43*)) iterations to generate a consensus tree, which formed the basis for the ancestral state reconstruction analysis.

To time-scale the consensus tree we used LSD2 v1.9.7 (*44*), using Wuhan-Hu-1 as an outgroup. We fixed the clock-rate to 5.6e4 as reported for B.1.1.7 previously (*12*) and collapsed branches with lengths < 1e5 to reduce noise. The -e option was set to four, removing sequences that deviated more than 4 standard deviations from the branch length distribution.

The time-scaled tree was used as input to pastml v1.9.33 (*21*) which we ran with default settings except setting the *--resolve_polytomies* option, as polytomies can inflate the number of introductions in the analysis (*45*). PastML was used to reconstruct ancestral states using a four state F81-like model for nucleotide substitution generalized to the number of geographic states (*46*). Under the F81-like model, the migration rate from a state $i$ (e.g. geographic region) to a different state $j$ ($i \neq j$) is proportional to the equilibrium frequency of $j$, termed $\pi j$. The rescaling factor which is analogous to the mutation rate under a strict molecular clock is optimized in PastML, in addition to the state equilibrium frequencies. In other words, the rescaling factor represents the average number of character changes per year for time-scaled trees. This is applied to all tree branches, which represents the average number of character changes per branch unit.

**List of Danish Covid-19 Genome Consortium members**

The ordering of affiliations and authors does not reflect any difference in contribution.

Kasper S. Andersen[1], Martin H. Andersen[1], Amalie Berg[1], Susanne R. Bielidt[1], Sebastian M. Dall[1], Erika Dvarionaite[1], Susan H. Hansen[1], Vibeke R. Jørgensen[1], Rasmus H. Kirkegaard[1], Wagma Saei[1], Trine B. Nicolajsen[1], Stine K. Østergaard[1], Rasmus F. Brøndum[2], Martin Bøgsted[2], Katja Hose[3], Tomer Sagi[3], Miroslaw Pakanec[3], David Fuglsang-Damgaard[4], Mette Mølvadgaard[4], Henrik Krarup[5], Christina W. Svarrer[6], Mette T. Christiansen[6], Anna C. Ingham[6], Thor B. Johannesen[6], Martín Basterrechea[6], Berit Lilje[6], Kirsten Ellegaard[6], Povilas Matusevicius[6], Lars B. Christoffersen[6], Man-Hung E. Tang[6], Kim L. Ng[6], Sofie M. Edslev[6], Sharmin Baig[6], Ole H. Larsen[7], Kristian A. Skipper[7], Søren Vang[7], Kurt J. Handberg[8], Marc T. K. Nielsen[8], Carl M. Kobel[8], Camilla Andersen[8], Irene H. Tarpgaard[8], Svend Ellermann-Eriksen[8], José A. S. Castruita[9], Uffe V. Schneider[9], Nana G. Jacobsen[9], Christian Ø. Andersen[9], Martin S. Pedersen[10], Kristian Schønning[10], Nikolai Kirkby[10], Lene Nielsen[11], Line L. Nilsson[11], Martin B. Friis[11], Thomas Sundelin[11], Thomas A. Hansen[11], Marianne N. Skov[12], Thomas V. Sydenham[12], Xiaohui C. Nielsen[13], Christian H. Schouw[13], Anders Jensen[14], Ea S. Marmolin[14], John E. Coia[15], Dorte T. Andersen[15]

[1] Department of Chemistry and Bioscience, Aalborg University.
[2] Department of Clinical Medicine, Aalborg University.
[3] Department of Computer Science, Aalborg University.
[4] Department of Clinical Microbiology, Aalborg University Hospital.
[5] Department of Molecular Diagnostics, Aalborg University Hospital.
[6] Statens Serum Institut.
[7] Department of Molecular Medicine (MOMA), Aarhus University Hospital.
[8] Department of Clinical Microbiology, Aarhus University Hospital.
[9] Department of Clinical Microbiology, Copenhagen University Hospital.
[10] Rigshospitalet, Copenhagen University Hospital.
[11] Department of Clinical Microbiology, Herlev Hospital.
[12] Odense University Hospital.
[13] Zealand University Hospital.
[14] Sygehus Lillebælt.
[15] Department of Clinical Microbiology, Sydvestjysk sygehus.

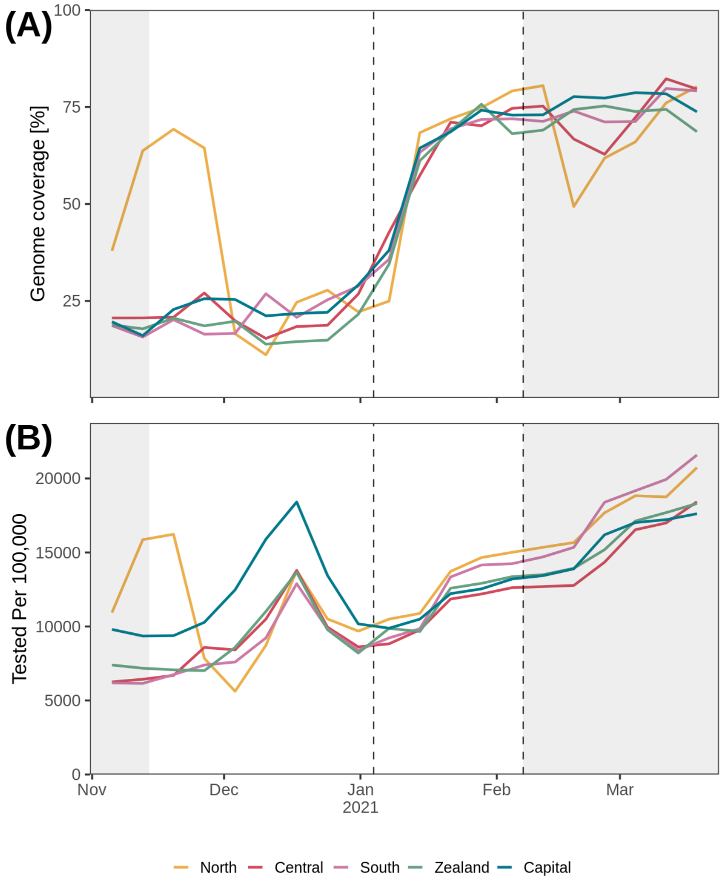


**Fig. S1.**

Sequencing rate relative to total number of covid19 cases per week **(A)** and relative testing effort **(B)** for each Danish region across time. The two vertical dashed lines indicate the beginning and end of study period used to infer B.1.1.7 transmissibility, while the non-shaded area shows the period used for phylogenetic analysis. The time outside the study are shaded in grey.

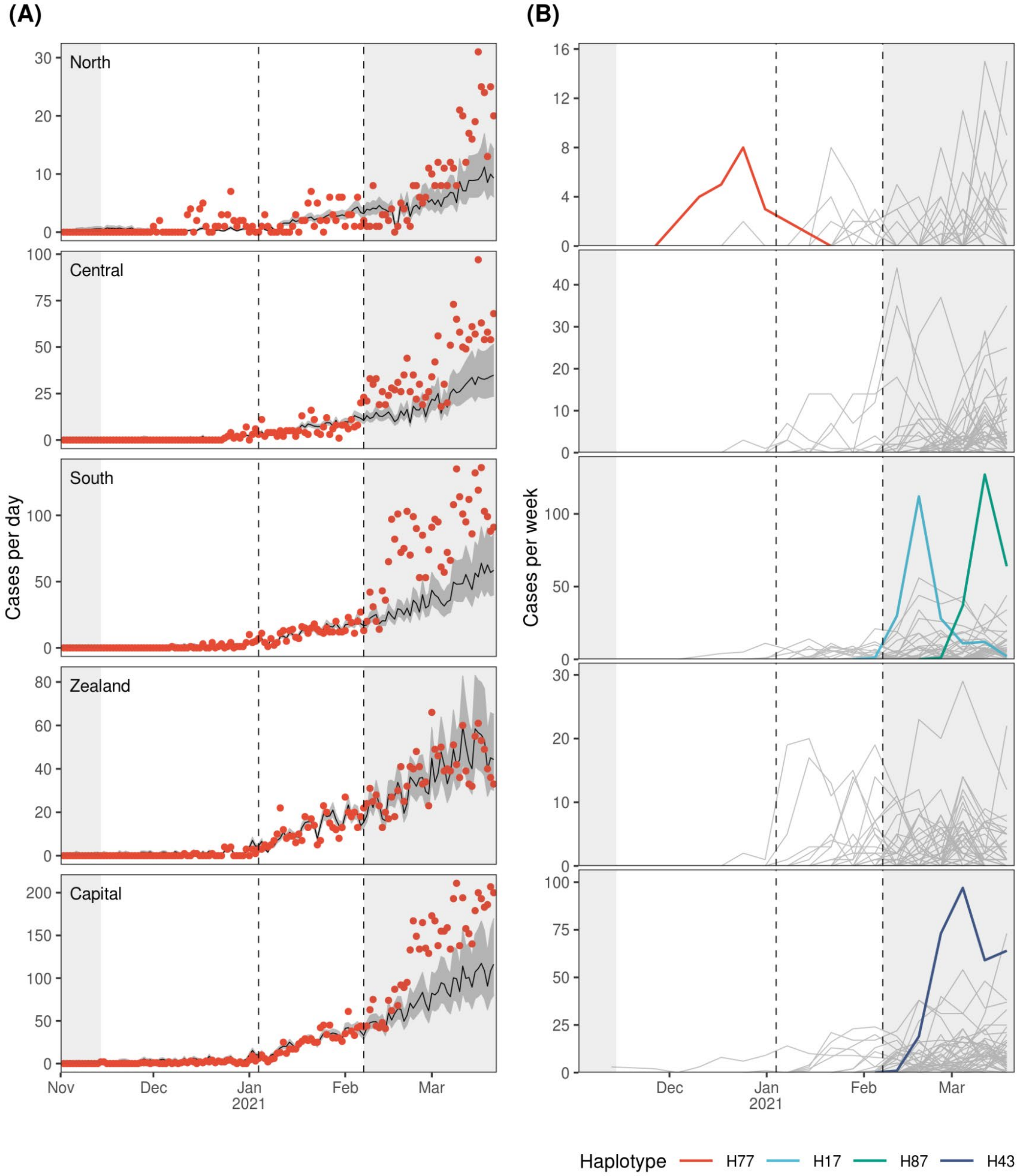


**Fig. S2.**

The two vertical dashed lines indicate the beginning and end of study period used to infer B.1.1.7 transmissibility, while the non-shaded area shows the period used for phylogenetic analysis. The time outside the study are shaded in grey. (**A**) Model predictions from Poisson regression model on daily counts of B.1.1.7 for each region. Dark-grey areas represent 95% CI. (**B**) Frequency of unique haplotypes across time for each region. Each line represents the weekly count of a unique B.1.1.7 haplotype. The four haplotypes mentioned in the main text are highlighted.

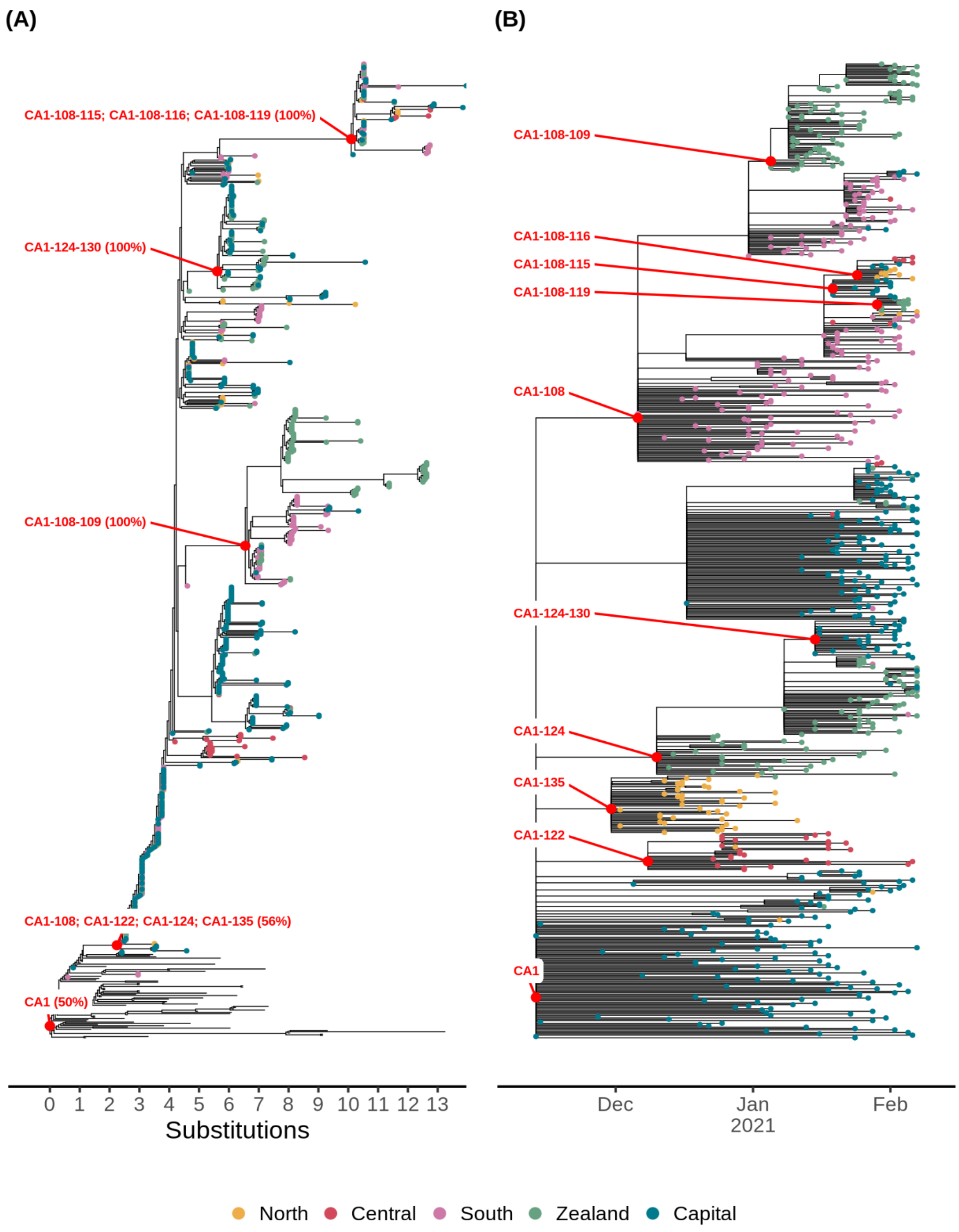


**Fig. S3.**

The phylogenetic structure of the data behind the introduction lineage CA1 and descendant transmission clusters as shown in fig. 2. Each point is coloured according to region, branches without points are sequences from outside Denmark. Red dots indicate the root node of each cluster with labels. (**A**) is the phylogenetic consensus tree generated from 1000 ultrafast-bootstrap iterations using the -B option in IQ-tree. Polytomy nodes are collapsed by ";" in the labels. Percentage values in each label indicate the bootstrap confidence. (**B**) time-scaled, pruned version of the tree in (**A**) used for ancestral state reconstruction.

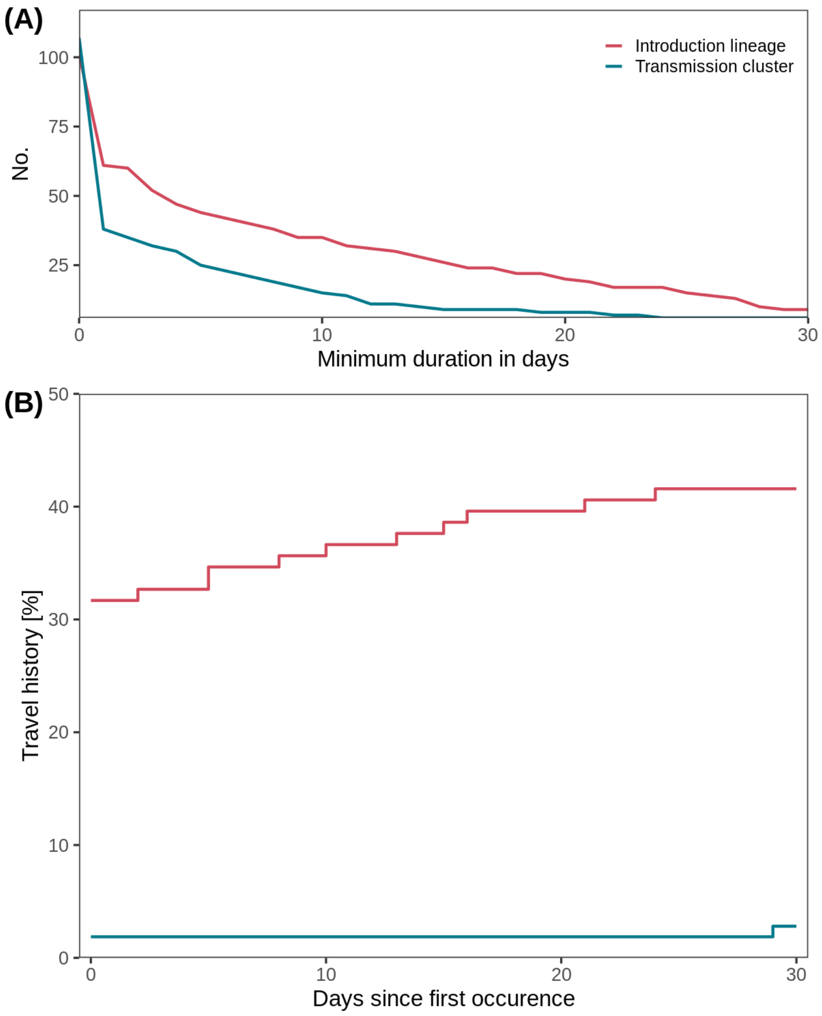


**Fig. S4.**

Assessing robustness of inferred introductions from phylogenetic analysis using travel history. (**A**) and (**B**) are grouped into introduction lineages that are introduced from abroad and transmission clusters introduced from other Danish regions. (**A**) shows the number of introduction lineages and transmission clusters with a minimum duration given on the x-axis. (**B**) shows the percent of introduction lineages and transmission clusters with travel-associated cases before a cutoff day indicated on the x-axis. The cutoff day on x-axis is relative to the first occurrence of the introduction lineage or transmission cluster.
